# An individual patient data (IPD) meta-analysis on negative effects of Mindfulness-Based Cognitive Therapy (MBCT) and Cognitive Behavior Analysis of Psychotherapy (CBASP) for patients with difficult-to-treat depression (DTD)

**DOI:** 10.1101/2025.06.09.25329260

**Authors:** Johannes Michalak, Maria Niemi, Maria Velana, Thorsten Barnhofer, Mathias Harrer

**Author notes:** Corresponding author: Associate Professor Maria Niemi, Department of Global Public Health, Karolinska Institutet, Stockholm, Sweden.

## Abstract

**Background:** Chronic and treatment-resistant depression remains a challenge. Although research has demonstrated positive effects of Mindfulness-Based Cognitive Therapy (MBCT) and Cognitive Behavioral Analysis System of Psychotherapy (CBASP), their potential negative effects—including deterioration, suicidality, and adverse events—are not well understood.

**Objectives:** This protocol outlines an individual patient data (IPD) meta-analysis to assess and compare the incidence and correlates of negative effects of MBCT and CBASP in adults with chronic and treatment-resistant forms of depression, subsumed under the wider heuristic of difficult-to-treat depression. Secondary aims include examining effects on depressive symptoms across these interventions and identifying moderators of negative outcomes.

**Methods:** Randomized controlled trials (RCTs) of MBCT and CBASP for adults (18–65 years) with chronic or treatment-resistant depression will be identified through systematic searches of major databases. Eligible studies must compare MBCT or CBASP (alone or with treatment as usual) to each other or to control groups. The primary outcome is clinically significant deterioration, defined as a ≥6-point increase on the PHQ-9 or equivalent. Secondary outcomes are suicidality, adverse events, and depressive symptom change. IPD will be requested from authors; aggregate data will be used if IPD is unavailable. One-stage random effects meta-analyses and moderator analyses will be conducted.

**Discussion:** This IPD meta-analysis will provide robust estimates of negative effects for MBCT and CBASP in DTD, informing clinical practice and guidelines on patient safety.

**PROSPERO registration number:** CRD42022332039

**Strengths and limitations:**

- Leveraging an individual patient data (IPD) meta-analysis allows for more precise assessment of negative effects and enables detailed moderator analyses that are not possible with aggregate data approaches.
- Standardizing outcome measures across studies using a common metric enhances comparability but may result in some loss of information or sensitivity to subtle differences in negative outcomes.
- Differences in the availability and reporting of individual-level variables across studies may limit the ability to fully explore all potential moderators of negative effects, and reliance on data sharing from original authors could restrict the inclusion of eligible studies.

## INTRODUCTION

According to the most recent Global Burden of Disease (GBD) study, the global prevalence of Major Depressive Disorder (MDD) is approximately 3.8% [1]. This figure highlights the significant impact of MDD on global health and underscores its status as a leading cause of disability worldwide. MDD is characterized by a broad range of symptom profiles that can range from mild to severe, exhibiting diverse responses to treatments and chronicity [2]. While establish treatments can help a significant number of patients, about a third enter a recurrent or chronic course in which established treatments fail to bring sustained remission [3]. Recently framed under the broader heuristic of difficult-to-treat depression (DTD), such courses are associated with significant continuing burden [3, 4].

Traditionally, pharmacological treatments have been the primary approach for managing DTD. However, there is a growing understanding that effective management should target a holistic approach that addresses symptoms, improves functionality, and reduces the overall burden [5]. Psychological therapies, which are favored by many patients, are essential in this context. Two such therapies are Mindfulness-Based Cognitive Therapy (MBCT) and the Cognitive Behavioral Analysis System of Psychotherapy (CBASP).

MBCT combines elements of cognitive therapy with mindfulness meditation and was originally introduced for the prevention of relapse in patients currently in remission [6]. It has been proven effective for this purpose, particularly in patients with highly recurrent courses [7]. Mindfulness involves cultivating a non-judgmental awareness of present-moment experiences, including thoughts, emotions, and bodily sensations. In the context of depression, this practice helps individuals disengage from negative thought patterns and rumination, which are common triggers for depression. More recent research has extended the use of MBCT to patients with current depression that has been proven difficult-to-treat [8-10]. Evidence suggests that MBCT can be beneficial for patients with DTD, warranting guideline endorsement and wider implementation [9, 10].

CBASP was specifically designed to treat chronic depression [11]. It focuses on interpersonal interactions and aims to alter maladaptive patterns of behavior through disciplined personal involvement with the therapist. Additional features of CBASP include the use of situational analysis to help patients understand the consequences of their behavior, the clarification of interpersonal goals, and the therapist’s use of techniques, such as “interpersonal discrimination exercises” to distinguish past from present relational experiences. The therapeutic relationship is central and is used as a tool to model healthy interactions and corrective emotional experiences [11]. Studies have shown that CBASP, especially when combined with antidepressant medication, can be effective for patients with chronic and treatment-resistant depression [11, 12].

Despite many studies demonstrating benefits of MBCT and CBASP for DTD, little attention has been given to their potential negative effects. As the field increasingly emphasizes not only the benefits, but also the risks of psychological treatments [13, 14], patient safety has become a critical concern. This individual participant data (IPD) meta-analysis aims to systematically investigate the incidence and correlates of negative effects associated with MBCT and CBASP, thereby contributing to a more nuanced understanding of their safety profiles.

Therefore, it is timely to bring together evidence from existing randomized controlled trials and analyze negative effects across studies of MBCT and CBASP for DTD. For this purpose, pooling IPD from these studies to conduct meta-analyses using one-stage random effects meta-analysis [15-17] is a suitable approach. This approach offers advantages over conventional aggregate data meta-analyses [15-17]. In line with this, IPD meta-analysis allows examination of moderator or subgroup questions across the pooled sample, overcoming limitations in sample size and power in the original studies. This information is crucial for implementation, providing information about factors that may increase or decrease risk of negative effects.

More specifically, the objectives of the present study are:

1. To compare the rates of clinically significant deterioration in MBCT, CBASP, and other controls in patients with difficult-to-treat depression
2. To compare levels of suicidality following MBCT, CBASP, and other controls for difficult-to-treat depression
3. To investigate and compare the rates of adverse events in MBCT, CBASP, and other controls in patients with difficult-to-treat depression

## METHODS

### Study design

This IPD meta-analysis will include any randomized controlled study investigating the efficacy of any two of MBCT, alone or adjunct to TAU, CBASP, alone or adjunct to TAU, and other controls (see “Comparator” section) for reducing symptoms of depression in eligible subjects. We will assess rates of clinically significant deterioration, suicidality and adverse events in MBCT, CBASP and other controls for DTD. This IPD meta-analysis is prospectively registered in PROSPERO and any key changes or amendments will be documented there. The Preferred Reporting Items for Systematic Reviews and Meta-Analyses IPD statement will be followed for the reporting of this study.

### Eligibility criteria

#### Condition or domain being studied

This meta-analysis will focus on the wider domain of DTD. As DTD does not link to an objective way of assessment, we will include studies that have used a range of conventional definitions encompassed by this wider heuristic, including studies that have selected participants based on the presence of chronic depression and treatment resistance in its broadest sense (including treatment non-response and non-remission).

#### Population

Treatment-resistant depression is defined broadly, including patients with non-response or non-remission following pharmacological and/or psychological treatment, encompassing one or more failed treatment attempts. Chronic depression will include adults meeting DSM-5 criteria for *persistent depressive disorder* or equivalent DSM-IV diagnoses, including dysthymic disorder, chronic major depression, current major depressive episode as part of a recurrent major depression with incomplete recovery between episodes during the last two years, and double depression (a current major depressive episode superimposed on antecedent dysthymia). We will also include adults with recurrent major depression who have experienced incomplete recovery between episodes, such that clinically significant depressive symptoms have been present continuously for at least two years. We will exclude adolescents (under 18 years of age) and adults older than 65.

#### Intervention(s) or exposure(s)

The interventions of interest will include manualized MBCT, delivered as a stand-alone treatment or as an adjunct to treatment as usual (TAU) [18]. MBCT is a structured psychotherapeutic intervention that incorporates elements of cognitive behavioral therapy and mindfulness-based stress reduction [19]. Mindfulness is commonly defined as non-judgmental awareness of the present moment [20]. As MBCT was originally designed for relapse prevention, we will accept studies that introduce minor changes to address acute depressive symptoms.

#### Comparator(s) or control(s)

In the network, MBCT will be compared to manualized CBASP, delivered alone or as an adjunct treatment to TAU [21], as well as to other eligible control conditions (e.g. TAU, waitlist, active psychological comparators, or other standard treatments). CBASP is psychotherapeutic intervention specifically developed for chronic depression, integrating elements of cognitive behavioral therapy and interpersonal/psychodynamic approaches [22].

#### Context

We will include studies that recruited patients in any setting.

#### Outcomes

The primary outcome will be rates of clinically significant deterioration measured using standard depression assessment tools, in self-report questionnaires (e.g., Patient Health Questionnaire 9, PHQ-9; Beck Depression Inventory) or observer-rated measures (Hamilton Depression Rating Scale). When both types of measures are available, we will analyze the effects on each tool separately. If multiple measures are used in a single study, preference will be given to the measure most commonly used across studies. To ensure comparability of symptom severity scores across different measures, values will be standardized using the common metric approach by Wahl et al.[23]. If a measure is not part of the common metric, it will be converted using validated conversion tables to derive the common metric. Common metric scores will be converted to the PHQ-9, and clinically significant deterioration and other dichotomous outcomes will be calculated based on this measure. According to PHQ-9 conventions, deterioration will be defined as an increase of 6 or more points.

#### Baseline constructs

Baseline constructs will include assessments of potential psychological moderators of adverse effects. These will include potential moderators at the level of the individual patient (e.g. sociodemographic and clinical characteristics) [24] and at the level of the study. We will include patient level characteristics in the analyses, if they are consistently available across the datasets and, where inclusion is justified, based on evidence of possible effects on treatment response from prior research.

Individual level sociodemographic and clinical characteristics that will be investigated include age, gender, marital status, education, employment, childhood adversity, age at onset of depression, number of previous episodes, current episode duration, current suicidality, current antidepressant medication, past antidepressants and treatment-resistance to antidepressants, co-morbid anxiety disorder, comorbid obsessive-compulsive disorder, co-morbid post-traumatic stress disorder, co-morbid substance abuse or dependence, co-morbid eating disorder, co-morbid personality disorder, and level of depressive symptoms at baseline [24]. Intervention and study characteristics will include number of treatment sessions attended, treatment completer status, type of delivery (remote versus face-to-face), and type of DTD (non-response/remission, treatment-resistance, chronic depression).

As an additional indicator of negative effects, we will analyze increases in suicidality by examining the specific suicidality items included in standard depression assessment tools. Furthermore, any reported adverse events and serious adverse events will be evaluated, provided that this information is available in the respective datasets

### Searches

We have searched the following databases for relevant publications: EMBASE, PubMed/MEDLINE, PsycINFO, Web of Science, Scopus, and the Cochrane Controlled Trials Register (see search terms in appendix 1). Once relevant publications have been identified, their bibliographic reference list will be searched for any additional relevant study. We have searched for any study published up until the day of the search, on May 30^th^ 2025 and there was no lower time limit. We searched for studies published in English and unpublished studies were not sought.

### Data extraction (selection and coding)

Following completion of the search strategies, initial screening of titles will be performed by two independent reviewers, with the publication abstract being obtained if selected by at least one reviewer. Abstracts will be screened by two reviewers, with disagreements about eligibility being referred to a third reviewer whose decision will be final. Records will be kept of all titles and abstracts screened, and reasons for decisions for inclusion and exclusion. Records will be signed by all three reviewers.

As this is an IPD meta-analysis, data will be sought, rather than extracted from publications. Corresponding authors responsible for the included studies will be contacted via email by the project leaders with an invitation to participate in the review and to share the IPD from their primary study. The standard letter will include all relevant details of the study including its purpose and main research questions, the variables of interest and will include a request to share the raw data from the study. Corresponding authors of studies will be invited to serve as collaborators to the project. If corresponding authors do not respond within four weeks, we will send a second email request. If this fails, a second author will be contacted. In case of further failure to make contact, we will continue until we have contacted a maximum of three authors. If none of the authors responds, or if the authors indicate that the data are unavailable, we will list the study data as unavailable. Each contacted author will receive an initial email and one reminder. If we are unable to get IPD for a study, we will extract aggregate data from the paper of this study.

### Risk of bias (quality) assessment

We will assess risk of bias for all included studies using the revised Cochrane Risk-of-Bias Tool for Randomized Trial (RoB2) tool [25, 26]. Each study will be assessed for quality by two independent reviewers, with disagreements referred to a third reviewer, whose decision will be final. A sensitivity analysis will be performed excluding studies judged to be at high risk of bias, or where risk of bias is unclear.

### Planned data synthesis

To calculate a pooled estimate of the (direct and indirect) treatment contrasts included in the obtained IPD, a one-stage IPD network meta-analysis (IPD-NMA) model will be employed, adjusting for baseline prognostic factors. The effect of moderators will be assessed by adding treatment-covariate interactions to this main IPD-NMA model on a one-by-one basis, yielding a IPD network meta-regression (IPD-NMR). In all models, covariates will be centred and scaled by cluster to facilitate convergence.

The primary outcome, symptom deterioration, will be modeled using a binomial logit-link. Let *y*_*ik*_ be the deterioration status of some participant *i* included in study *k* (with *k*=1, …, *K* studies included the meta-analysis), where *j=b* represents the baseline arm, and *j=t* the intervention arm in *k*. The one-stage IPD-NMA model will then be defined as:

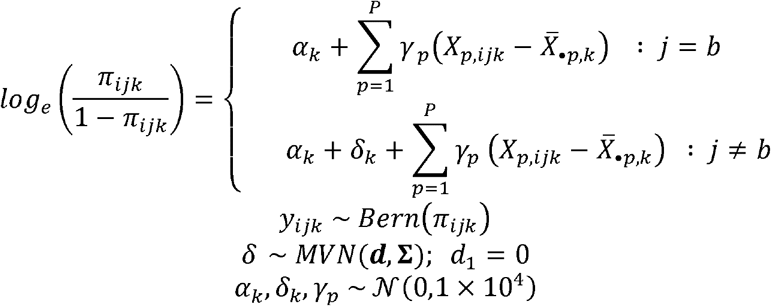

where ***d*** represents a vector containing the effect sizes in comparison to the reference treatment implicated by our network, and with ∑ being a heterogeneity variance-covariance matrix with off-diagonal elements *ρ τ*^2^, *ρ* where is set to a non-zero value to account for multi-arm studies. A weakly informative *ℋC*(0,5)prior will be used by default for the square root of the heterogeneity variance, *τ*.

The presence of network consistency will be explored a posteriori by calculating the agreement between direct and indirect evidence for the MBCT-CBASP comparison in an inconsistency model including an additional inconsistency parameter *w*.

As a sensitivity analysis, we will (1) perform pairwise one-stage IPD meta-analyses using the direct evidence in the network and (2) employ two-stage IPD-NMA models additionally.

All models will be fitted within a Bayesian framework using Gibbs sampling as implemented in JAGS [27]. The potential scale reduction factor (PRSF; 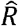) will be used to assess convergence. Flat normal or weakly informative half-Cauchy priors will be used for all model parameters. Missing data will be handled using multilevel multiple imputation with *m*=50 sets. All models will be fitted within the multiply imputed data, and parameter estimates will be aggregated by mixing the posterior draws.

### Analysis of subgroups or subsets

We will use the IPD dataset to investigate differential treatment effects across participant characteristics, such as baseline scores of the outcome variable and a history of early trauma, by pooling the stratified treatment effect modification coefficients of relevant predictors.

### Patient and public involvement

A lived experience advisory group will be consulted to provide input on the interpretation of the study findings and the development of dissemination materials. The results will be shared in accessible formats, including a lay summary co-produced with individuals with lived experience, to ensure findings are communicated effectively to patients and the wider public.

## DISCUSSION

This IPD meta-analysis will address a critical yet often underexplored aspect of psychotherapeutic treatment: the occurrence of negative effects, including symptom deterioration, suicidality, and adverse events. While there is encouraging evidence for the efficacy of MBCT and CBASP across the wider spectrum of DTD, potential harms remain insufficiently studied. By pooling individual-level data across trials, this study will provide a more precise and nuanced understanding of the prevalence, severity, and predictors of negative outcomes in these therapies.

One of the strengths of this study is the use of IPD, which enables detailed subgroup analyses and the examination of individual-level moderators. Inclusion of participant-level prognostic factors may also help to improve the plausibility of the transitivity assumption, which is crucial for the interpretability of NMA models – this is an advantage of analyzing IPD versus aggregate data alone. However, there are potential limitations, including the variability in study designs and the number and/or quality of the included trials. Additionally, the transformation of different outcome measures into a common metric may result in some loss of information and/or measurement error. Despite these limitations, the findings of this analysis have the potential to inform both clinical decision-making and the development of guidelines that better balance benefit and risk. A clearer understanding of which patients are most vulnerable to negative effects may also support more personalized treatment approaches and enhance informed consent procedures.

## Data Availability

This is individual participant data from randomized controlled trials, which cannot be shared publicly due to ethical and consent restrictions. Interested parties will be asked to contact the corresponding author to request data access. Access may be granted if the following conditions are met:(1) all co-authors approve for the data sharing, (2) a data-sharing agreement is in place, (3) individual studies have participant consent and ethics approvals in place to allow for further onward sharing, and (4) an analysis plan has been developed and agreed upon by all co-authors. Once shared, the data may only be used for the specified purposes outlined in the agreement.

## Ethics and Dissemination

No local ethical review was necessary following consultation with the Swedish Ethical Review Authority. The investigators of the primary trials have obtained local ethical approval for sharing their data. TB and MN will oversee the database, and data analyses will be conducted by MH. Patient privacy will be ensured by adhering to the Karolinska Institutet guidelines on data management and storage. Data exchange is governed by inter-institutional data sharing agreements, and all data were pseudonymized before sharing. The data set will not be open access, but access may be shared upon reasonable request.

The results of this meta-analysis will be disseminated through peer-reviewed journals, lay summaries, and discussions with patient and lived experience groups. Additional dissemination methods will include conference presentations and social media announcements. By making the findings accessible to various stakeholders, we aim to enhance the understanding and implementation of MBCT and CBASP in treating DTD.

## Contributions

JM, TB, MN, MV and MH conceptualized and designed the study. JM, TB and MN developed the search strategy. JM, TB, MN and MV contacted the primary authors, who contributed to further refinement of the design and approach of the study. TB, MV and MN are responsible for building the database. MH is responsible for data analyses and will provide statistical expertise. MN drafted the protocol manuscript, which was critically revised by all authors. All authors read and approved the final version. JM and MN serve as guarantors of the review.

## Competing interests statement

TB is the co-author of a book on mindfulness-based interventions and regularly offers workshops on mindfulness-based interventions. He is the co-investigator of a programme grant evaluating an adapted MBCT course for adolescents experiencing depression.

MN is the author of a book chapter om mindfulness-based interventions and offers workshops on mindfulness-based interventions.

MV reports no conflicts of interest.

JM is the director of the Achtsamkeitsinstitut Ruhr (an institute offering mindfulness training) and principal investigator of several DFG (German Science Foundation) research projects. He receives royalties from mindfulness books he has authored.

In the past three years, MH served as a statistical consultant for HelloBetter/GetOn Institut fu□r Gesundheitstrainings GmbH, a company that implements digital mental health therapeutics in routine care.

## Appendix 1: Search Terms for CBASP

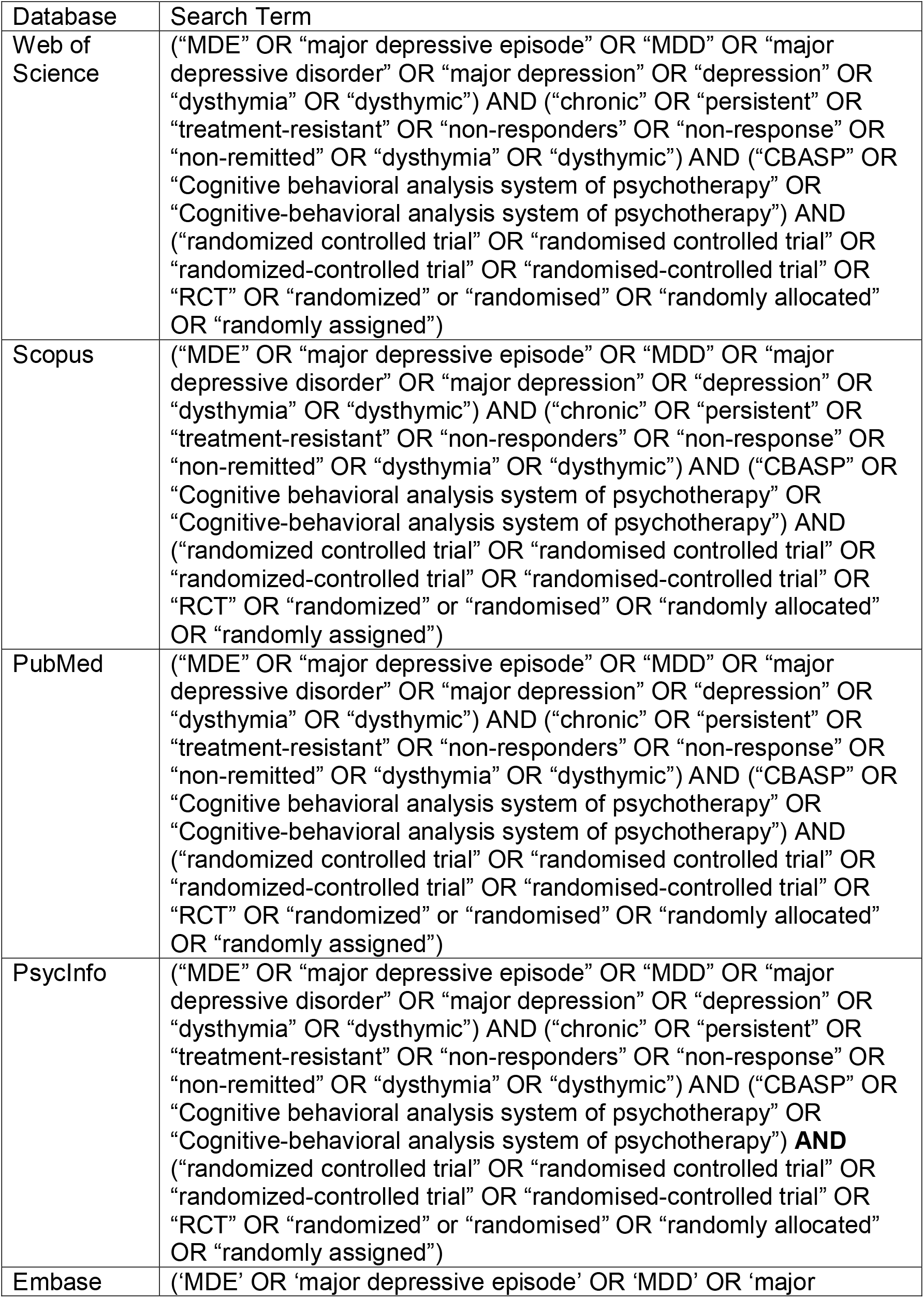
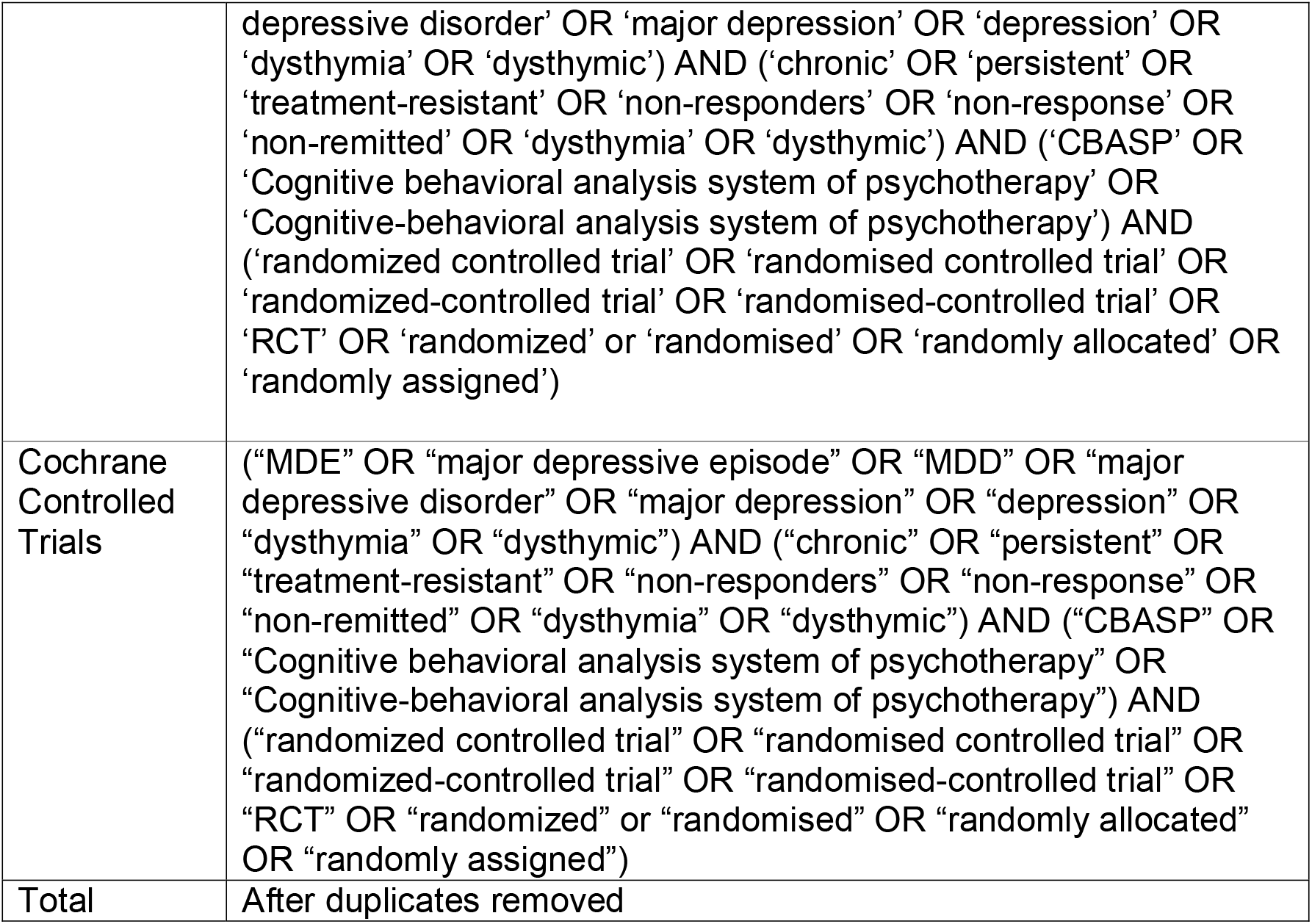

### Search Terms for MBCT

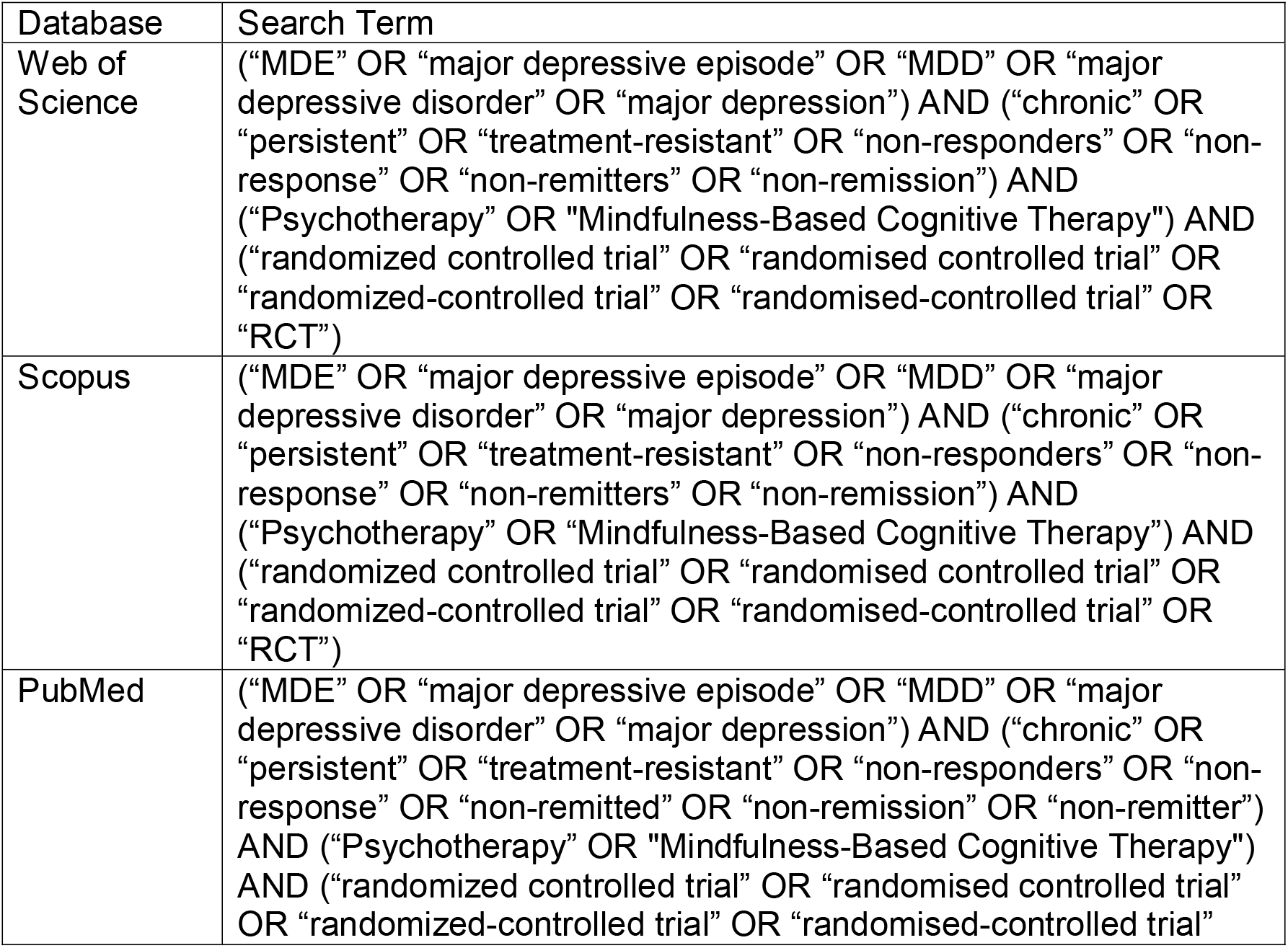
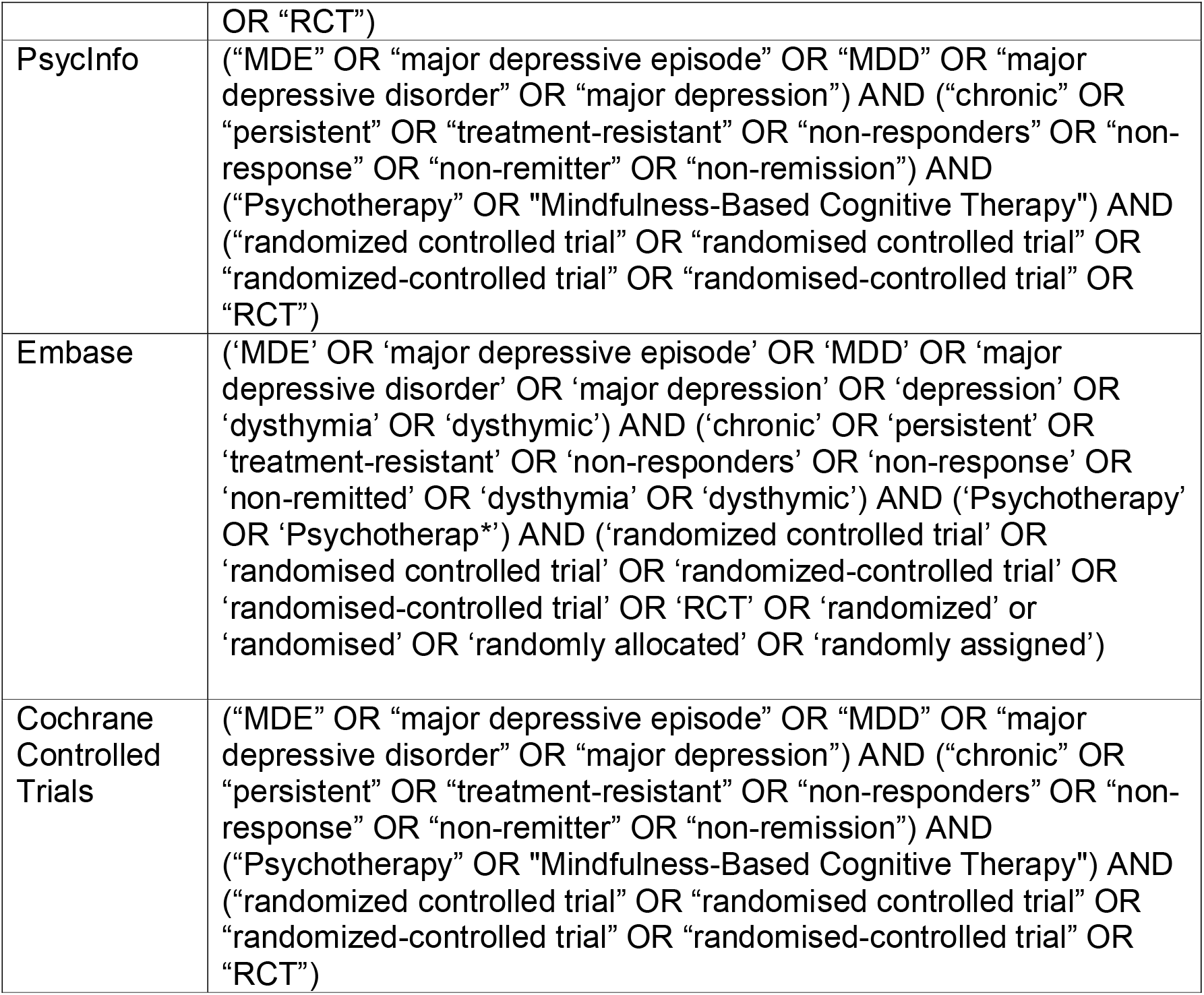

